# Accuracy of tongue swab testing using Xpert MTB-RIF Ultra for tuberculosis diagnosis

**DOI:** 10.1101/2022.02.17.22271147

**Authors:** A Andama, GR Whitman, R Crowder, TF Reza, D Jaganath, J Mulondo, TK Nalugwa, C. F Semitala, W Worodria, C Cook, RC Wood, KM Weigel, AM Olson, J Lohmiller Shaw, C Denkinger, P Nahid, G Cangelosi, A Cattamanchi

**Affiliations:** Department of Medicine, College of Health Sciences, Makerere University, Kampala, Uganda; Infectious Diseases Research Collaboration, Kampala, Uganda; Department of Environmental and Occupational Health Sciences, School of Public Health, University of Washington, Seattle, WA USA; Uganda Tuberculosis Implementation Research consortium, Kampala, Uganda; Division of Pulmonary and Critical Care Medicine and Center for Tuberculosis, San Francisco General Hospital, University of California, San Francisco, CA USA; Division of Pediatric Infectious Diseases, University of California, San Francisco, CA USA; Mulago National Referral Hospital, Kampala, Uganda; University Hospital of Heidelberg, Germany

## Abstract

Tongue dorsum swabs have shown promise as alternatives to sputum for detecting *Mycobacterium tuberculosis* (MTB) in patients with pulmonary tuberculosis (TB). Some of the most encouraging results have come from studies that used manual quantitative PCR (qPCR) to analyze swabs. Studies using the automated Cepheid Xpert® MTB/RIF Ultra qPCR test (Xpert Ultra) have yielded less encouraging results with tongue swabs, possibly because Xpert Ultra is optimized for testing sputum, not tongue swab samples. Using two new sample processing methods that demonstrated good sensitivity in preliminary experiments, we assessed diagnostic accuracy and semi-quantitative signals of 183 tongue swab samples using Xpert Ultra in a clinical setting. Relative to a sputum Xpert Ultra reference standard, the sensitivity of tongue swab Xpert Ultra was 68.5% (95% CI 54.4-80.5) and specificity was 100.0% (95% CI 97.2-100.0). When compared to a microbiological reference standard (MRS) incorporating both sputum Xpert Ultra and sputum culture, sensitivity was 64.9 (95% CI 51.1-77.1) and specificity remained the same. Higher sensitivity was observed (77.8% CI 64.4-88.0) when “trace” Xpert results were included among positive swabs. Semi-quantitative Xpert Ultra results were generally lower with swabs than with sputum. None of the eight sputum Xpert Ultra “trace” or “very low” results were detected using tongue swabs. Tongue swabs should be considered when sputum cannot be collected for Xpert Ultra testing, or in certain mass-screening settings. Further optimization of tongue swab analysis is needed to achieve parity with sputum-based molecular testing for TB.

## 1. Introduction

Each year, an estimated 10 million people develop tuberculosis (TB) and over one million die of TB despite the availability of effective treatment for most forms of the disease [1]. A significant barrier to current TB care and control efforts is the difficulty of rapidly screening large numbers of people for active disease. Historically, TB diagnosis has relied on passive case finding, in which people with symptoms self-report to a healthcare facility for further evaluation. Pulmonary TB diagnosis most often relies on microbiological or molecular analysis of sputum for the presence of *Mycobacterium tuberculosis* (MTB) cells or DNA to confirm diagnosis [2]. The production of sputum can be burdensome for those providing the specimen and hazardous to health workers and others when adequate safety measures are not in place. Moreover, children and people living with HIV (PLHIV) are often unable to produce adequate sputum [3, 4]. These limitations have spurred efforts to identify alternative sample types for TB detection that are easier, less invasive, and safer to collect.

Molecular analysis of sputum, such as with Xpert MTB/RIF Ultra (Xpert, Cepheid, USA) is now the first-line test recommended for diagnosis of pulmonary TB [5-7]. Sputum sample processing for Xpert testing involves the addition of Cepheid’s proprietary Sample Reagent (SR). This reagent liquefies and decontaminates the sample without lysing bacilli. Once the treated sample is loaded into the cartridge and the run started, the sample is drawn through a membrane filter, trapping intact bacilli. After wash steps, a sonic horn lyses the bacilli on the membrane and releases their DNA. The DNA is then subjected to a semi-nested quantitative polymerase chain-reaction (qPCR), targeting the rifampicin resistance determining region (RRDR) of the rpoB gene. A mixed two-insertion sequence (IS1081 and IS6110) probe is also included to enhance detection of low bacillary load samples [5, 8].

Tongue swabbing, also known as oral swab analysis (OSA), has emerged as a potential alternative to sputum-based molecular testing [9]. Compared to sputum collection, tongue swabbing is faster, easier, and safer. Because tongue swabs are easy to collect from any person in any setting, OSA may be especially useful for non-clinical and community-based screening. We have previously optimized and clinically validated OSA procedures for manual DNA extraction and IS6110-targeted qPCR [10-14]. Sensitivity in adults relative to positive sputum Xpert or positive sputum culture has ranged from 88% to 93%, with specificity ranging from 79% to 92% [11, 13].

Given the widespread use of Xpert for TB diagnosis in high burden countries, it is important to harmonize protocols for Xpert testing using new sampling methods. To date, only a few studies have evaluated OSA in conjunction with Xpert testing. Success has been limited, with reported sensitivities ranging from ∼45% relative to sputum culture [15] to ∼51% relative to sputum Xpert [16]. Although these studies utilized different swabbing and sample handling methods, the results underscore the need to optimize and standardize Xpert protocols for detecting MTB in tongue swabs.

In this study, we sought to identify and characterize improved methods and provide updated guidance and protocols for use of Xpert to detect MTB in tongue swabs using tongue swab samples spiked with standardized cultured MTB cells. We then evaluated the diagnostic accuracy of tongue swab Xpert when using the updated protocols among adults being evaluated for pulmonary TB at two outpatient clinics in Kampala, Uganda.

## 2. Materials and Methods

### 2.1. Method optimization experiments

Contrived samples consisting of tongue swabs collected from healthy volunteers spiked with cultured MTB were used for optimization of tongue swab processing methods (Supplemental information). Study participants for these experiments were adults (>18 years) recruited from the University of Washington, School of Public Health, Department of Environmental and Occupational Health Sciences. Participants provided samples between November 2019 and February 2021. Participants were assumed to be TB-negative based on low risk of exposure and lack of symptoms. Inclusion criteria and screening protocols have been described previously by Wood et al, 2015 [12]. A total of 15 participants were enrolled and sampled. Participants were contacted for repeated sampling as needed. In order to protect the safety of participants and study personnel, swabs were self-collected with study staff leaving the room while collection took place. Procedures for collection of swab samples from human participants were approved of by the University of Washington Human Subjects Division (STUDY00001840).

### 2.2. Clinical evaluation

For the clinical evaluation of Xpert performed on tongue swabs, we conducted a cross-sectional study between April and September 2021 at outpatient clinics at Mulago National Referral Hospital and Kisenyi Health Center IV in Kampala, Uganda. We screened consecutive adults (>18 years) presenting to the health centers for any reason for cough lasting more than two weeks. Among those with cough lasting more than two weeks, we excluded patients who 1) had completed latent or active TB treatment within the past 12 months; 2) had taken any medication with anti-mycobacterial activity (including fluoroquinolones) for any reason, within 2 weeks of study entry; 3) resided >20km from the study site or were unwilling to return for follow-up visits; or 4) were unwilling or unable to provide informed consent. The Makerere University School of Medicine Research and Ethics Committee, the Uganda National Council for Science and Technology and the University of California San Francisco Committee on Human Research reviewed and approved the study. The study was conducted and reported in accordance with the Standards for Reporting of Diagnostic Accuracy Studies (STARD) guidelines [2].

### 2.3. Method development and limit of detection (LoD) quantification

Using Copan FLOQSwab® tongue swabs collected from TB-negative volunteers and spiked with cultured MTB cells, we evaluated three strategies for improving the sensitivity of Xpert performed on tongue swabs. The LoD of each method was compared to that of manual methods reported previously [11, 13]. Methods are presented in detail in the Supplemental Information and summarized here. In Method 1, a single FLOQSwab was processed with Cepheid Sample Reagent (SR) using a protocol similar to that recommended by the manufacturer for sputum. In Method 2, two FLOQSwabs were collected and combined into a single tube and processed as in Method 1. In Method 3, a single FLOQSwab was processed by boiling, incubation, and mixing (without using SR). Methods 1 and 2, which used SR, were assessed because they closely resemble sputum processing protocols currently recommended by Cepheid for Xpert Ultra. Method 2 (SR, two swabs) tested the hypothesis that the processing steps applied to one FLOQSwab in Method 1 could be applied to two FLOQSwabs to improve the sensitivity of TB detection. A recent study demonstrated that a single flocked swab collects ≤10% of the tongue dorsum biomass that is available for TB testing [13]. Therefore, if two flocked swabs are collected in succession, suspended in buffer at the same volume as a single swab, and tested as a single sample, then the collection of MTB bacilli from TB patients may be doubled on average relative to single swabbing. Method 3 (boil method) was selected for this study because it resembled the first steps of our previous manual methods [11-14].

Preliminary evaluations of these methods used tongue swab samples collected from healthy US volunteers as follows. Samples were collected using COPAN FLOQSwabs® Regular Flocked Swab with an 80 mm or 30 mm breakpoint (Copan Italia SpA, Brescia Italy). These swabs were shown to maximize tongue dorsum biomass collection, relative to other swab products [13]. Participants self-swabbed along the breadth of the mid-tongue dorsum, firmly pressing and rolling the swab head for approximately 10 seconds. Swab heads were then immediately spiked with 10 µL of serially-diluted cultured MTB strain H37Ra stored in 1x phosphate buffered saline with 15% glycerol and 0.05% Tween® 80 (PBSGT), or with blank PBSGT. Swab heads were then broken off into 5 mL polypropylene transport tubes (Corning) containing 800µL sterile 1x Tris-EDTA (TE) buffer (10 mM Tris-HCl containing 1 mM EDTA•Na_2_, pH 8.0) (Corning) for Methods 1-3.

LoD’s of the experimental methods were quantified using serial dilutions as described in the Supplemental Information. LoDs are reported on a colony forming unit (CFU) per swab basis (which is the same as the CFU per sample amount for Xpert Methods 1 and 3, but is half of the CFU per swab basis for Method 2 which included 2 swabs each spiked with the same number of bacilli as a single swab). Raw dose-response data are shown in **Table S1**. Exploratory runs at some dilutions (*e.g*., 100 cfu/swab) increased the number of runs at those dilutions, relative to other dilutions. Results of all experiments were used in LoD calculations. This minimized bias that could have resulted from the designation of specific runs as exploratory.

### 2.4. Clinical evaluation procedures

Upon enrollment at the outpatient clinics, we obtained detailed demographic and medical history using a standardized form, collected blood for HIV testing and CD4 count (if HIV-positive) and collected up to three expectorated sputum specimens. One sputum sample was used to perform Xpert testing for all patients (an additional sample was collected and tested if the initial Xpert result was trace-positive). If the initial or repeat sputum Xpert was negative or invalid, the remaining two sputum samples were cultured in liquid media. Trained laboratory technologists conducted all TB testing following standard protocols for Xpert testing [17] and mycobacterial culture [18].

For culture, sputum samples were digested and decontaminated using the sodium hydroxide/N-acetyl-L-cysteine method with a final concentration of 1.5%, neutralized with sterile phosphate-buffered solution, centrifuged, and re-suspended in phosphate-buffered solution. One Mycobacterial Growth Indicator Tube (MGIT) was inoculated with each 0.5mL decontaminated sample. When MGIT tubes were not available, solid 7h10 media was used for culture, and in the event of a negative 7h10 culture result, leftover sputum was preserved for culture in liquid media (MGIT) when available. During a period where MGIT tubes were not available, 32 patients had mycobacterial cultures performed using 7H10 media (of whom 28 subsequently had MGIT cultures done using frozen sputum sediment).

Prior to inoculation a cocktail of antibiotics containing PANTA (polymyxin, amphotericin B, nalidixic acid, trimethoprim, azlocillin) mixed with OADC (oleic acid, bovine serum albumin, dextrose, and catalase) were added to MGIT tubes. MGIT tubes were incubated in a BACTEC MGIT 960 instrument (BD; Franklin Lakes, USA) for up to 42 days. Blood agar plates were prepared to check for contamination of positive MGIT tubes, and a stained Ziehl Neelsen (ZN) smear was examined to check for the presence of acid fast bacilli (AFB). Speciation was then performed to confirm the presence of *M. tuberculosis* complex using SD Bioline strips (SD MPT64TB Ag kit, South Korea).

Prior to sputum collection, patients were also asked to self-collect up to 3 tongue swabs (COPAN FLOQswab) using the same procedure described for healthy volunteers in Washington. Two tongue swabs were placed in a single tube with 800µL of TE buffer and processed within one hour for Xpert testing using Method 2. In a subset of patients, an additional single swab stored at −80 degrees Celsius in a tube with 800µL of TE buffer was processed for Xpert testing using Method 3.

### 2.5. Definitions of reference standard and index tests for the clinical evaluation

We considered patients to have TB if they had a positive result on sputum Xpert Ultra (grade very low, low, medium, high), two trace-positive sputum Xpert Ultra results or a culture result positive for MTB. We considered patients not to have TB if they had no positive result (sputum Xpert Ultra and culture), and two negative cultures. We considered patients to have indeterminate TB status if they had no positive result (sputum Xpert Ultra and culture) or <2 trace-positive sputum Xpert Ultra results but <2 negative MGIT cultures due to contamination.

Staff performing tongue swab Xpert Ultra (the index test) were blinded to reference standard TB test results. The primary clinical evaluation analysis for Methods 2 and 3 followed the WHO recommendations for interpreting Xpert results, with trace-positive results counted as positive for HIV+ participants only. Secondary analyses considered trace-positive results positive for all participants.

### 2.6. Statistical approach of the clinical evaluation

We summarized demographic and clinical data using appropriate descriptive statistics. We calculated sensitivity and specificity of tongue swab Xpert in reference to sputum Xpert results alone, and in reference to sputum Xpert and culture results (with TB status defined as described above). The difference in sensitivity between Method 2 and Method 3 was assessed using McNemar’s test. We used STATA 15 (StataCorp, College Station, TX, USA) for all analyses.

## 3. Results

### 3.1 Limits of detection of tongue swab processing methods for Xpert testing

**Figure S1** plots the limits of detection (LoD) curves of each of the described methods, and LoDs are summarized in Table 1. Method 1 (SR, single swab), with 69 spiked samples, was relatively insensitive, with an LoD of 101.7 CFU/swab (95% CI 64.5 – 144.0). With 64 spiked samples, Method 2 (SR, double swab) improved sensitivity with a LoD of 76.5 CFU/swab (95% CI 54.2 – 104.1). Method 3 (boil method) demonstrated the highest sensitivity, equal to or better than the manual methods used previously (Table 1). With a total of 38 spiked samples represented in the dose-response data, Method 3 exhibited an LoD of 22.3 CFU/swab (95% CI 15.3 – 34.3). For purposes of comparison, with 40 spiked samples, the LoD of our clinically-validated manual qPCR method was 53.5 CFU/swab (95% CI 36.9 – 73.0). This value fell between those of Methods 1 and 2 (**Table 1**), and therefore Method 2 was selected for the main clinical evaluation. Of note, Methods 1-3 had no false positives among negative control tongue swab samples (n = 27). Additionally, there were no false-positive rifampin resistance determinations (n = 61).

**Table 1.** Comparison of Methods 1-3 and manual qPCR method LoDs.

| Method | Description | H37Ra LoDs in cfu/swab (95% CI)<sup>1</sup> |
| --- | --- | --- |
| Method 1 | 1 FLOQswab, SR<sup>2</sup>, Xpert Ultra (“single swab SR”) | 101.7 (64.5 - 144.0) |
| Method 2 | 2 FLOQswabs, SR, Xpert Ultra (“double swab SR”) | 76.5 (54.2 - 104.9) |
| Method 3 | 1 FLOQswab, boil w/o SR<sup>2</sup>, Xpert Ultra (“boil method”) | 22.3 (15.3 - 34.3) |
| Manual (Reference)<sup>3</sup> | 1 FLOQswab, Qiagen extraction and EtOH precipitation, manual IS6110 qPCR | 47.6 (32.1 – 65.8) |

<sup>1</sup>LoDs, limits of detection. Contrived samples were tongue swabs from healthy volunteers spiked with dilution series of cultured MTB H37Ra.
<sup>2</sup>SR, GeneXpert Sample Reagent
<sup>3</sup>Method used in Luabeya et al (2019) and Wood et al (2021)

### 3.2. Study population

Between April 14 and September 9, 2021, 184 eligible patients were enrolled and underwent tongue swab Xpert testing using Method 2 (SR, two swabs). One patient had an indeterminate tongue swab Xpert result and was excluded from this analysis. Among the remaining 183 patients, median age was 33 years; 76 (41.5%) were female, 58 (31.7%) were HIV-positive, and 22 (12.0) had previously been diagnosed with TB (**Table 2**). TB symptoms were common, with 142 (77.6%) experiencing weight loss, 137 (74.9%) fever, 122 (66.7%) night sweats, 115 (62.8%) decrease in appetite, 26 (14.2%) bumps in neck, armpit, or groin, and 25 (13.7%) had hemoptysis (**Table 2**).

**Table 2.** Demographic characteristics by Method 2 tongue swab Xpert Ultra result.

|  | Total<br>(N=183) | Tongue swab<br>Ultra* Negative<br>(n=146) | Tongue swab<br>Ultra* Positive<br>(n=37) |
| --- | --- | --- | --- |
| **Female** | 76 (41.5) | 65 (44.5) | 11 (29.7) |
| **Age, median (IRQ)** | 33 (26-43) | 35 (28-45) | 26 (23-35) |
| **HIV-positive** | 58 (31.7) | 53 (36.3) | 5 (13.5) |
| **CD4 count<100** | 10 (17.2) | 9 (17.0) | 1 (20.0) |
| **Prior TB** | 22 (12.0) | 17 (11.6) | 5 (13.5) |
| **Smoked tobacco in last 7 days** | 16 (8.7) | 11 (7.5) | 5 (13.5) |
| **BCG** | 158 (86.3) | 128 (87.7) | 30 (81.1) |
| **BMI, median (IQR)** | 21.4 (19.3-23.9) | 21.7 (19.8-24.6) | 19.6 (17.9-20.9) |
| **Underweight (BMI<18.5)** | 36 (19.7) | 23 (15.8) | 13 (19.7) |
| **Symptoms** |  |  |  |
| **Cough >2 weeks** | 183 (100) | 146 (100) | 37 (100) |
| **Fever** | 137 (74.9) | 103 (70.6) | 34 (91.9) |
| **Hemoptysis** | 25 (13.7) | 17 (11.6) | 8 (21.6) |
| **Night sweats** | 122 (66.7) | 97 (66.4) | 25 (67.6) |
| **Weight loss** | 142 (77.6) | 108 (74.0) | 34 (91.9) |
| **Decrease in appetite** | 115 (62.8) | 86 (58.9) | 29 (78.4) |
| **Bumps in neck, armpit, groin** | 26 (14.2) | 20 (13.7) | 6 (16.2) |
| **Proteinuria 2+** | 21 (11.5) | 19 (13.0) | 2 (5.4) |

TB: tuberculosis
\* Tongue swab Ultra conducted using Method 2 (SR, two swabs). Trace results considered positive for HIV+ participants only.

### 3.3. Diagnostic accuracy

Relative to sputum Xpert, the sensitivity of tongue swab Xpert was 68.5% (95% CI 54.4, 80.5) and specificity was 100.0% (95% CI 97.2-100.0). Sensitivity reduced to 64.9% (95% CI 51.1, 77.1) against the microbiological reference standard and specificity remained 100% (95% CI 96.8, 100). When all trace tongue swab Xpert results were considered positive, sensitivity improved to 77.8% (95% CI 64.4, 88.0) with no impact on specificity (**Table 3**). Sensitivity of tongue swab Xpert was consistently higher among patients with a higher mycobacterial load, as assessed using sputum Xpert Ultra (**Table 4**). Semi-quantitative Xpert results were generally lower with tongue swabs than with sputum samples, with none of the eight sputum Xpert “trace” or “very low” results being detected by tongue swabs (**Table 5**).

**Table 3.** Diagnostic accuracy of tongue swab Xpert Ultra.

| **Tongue swab Ultra, Method 2 (N=183)**<br>**trace+ for HIV+ only** |  |  |
| --- | --- | --- |
|  | **Sputum Xpert** | **Microbiologic reference standard\*** |
| **Sensitivity** | 68.5 (54.4-80.5) | 64.9 (51.1-77.1) |
| **Specificity** | 100 (97.2-100) | 100 (95.8-100) |
| **PPV** | 100 (90.5-100) | 100 (90.5-100) |
| **NPV** | 88.4 (82.0-93.1) | 81.3 (72.6-88.2) |
| **Tongue swab Ultra, Method 2 (N=183)**<br>**trace+ for all** |  |  |
|  | **Sputum Xpert** | **Microbiologic reference standard\*** |
| **Sensitivity** | 77.8 (64.4-88.0) | 73.4 (59.1-83.3) |
| **Specificity** | 100 (97.2-100) | 100 (96.9-100) |
| **PPV** | 100 (91.6-100) | 100 (91.6-100) |
| **NPV** | 91.5 (85.6-95.5) | 88.1 (81.5-93.1) |
| **Tongue swab Ultra, Method 3 (N=37)**<br>**trace+for HIV+ only** |  |  |
|  | **Sputum Xpert** | **Microbiologic reference standard\*** |
| **Sensitivity** | 62.9 (44.9-78.5) | 62.9 (44.9-78.5) |
| **Specificity** | 100 (15.8-100) | 100 (15.8-100) |
| **PPV** | 100 (84.6-100) | 100 (84.6-100) |
| **NPV** | 13.3 (1.7-40.5) | 13.3 (1.7-40.5) |
| **Tongue swab Ultra, Method 3 (N=37)**<br>**trace+ for all** |  |  |
|  | **Sputum Xpert** | **Microbiologic reference standard\*** |
| **Sensitivity** | 62.9 (44.9-78.5) | 77.1 (59.9-89.6) |
| **Specificity** | 100 (15.8-100) | 100 (15.8-100) |
| **PPV** | 100 (84.6-100) | 100 (87.2-100) |
| **NPV** | 20.0 (2.5-55.6) | 20.0 (2.5-55.6) |

\* excluding 6 indeterminate MRS results (<2 negative cultures due to contamination)

**Table 4.** Sensitivity of tongue swab Xpert Ultra compared by sputum Xpert Ultra semiquantitative grace.

| Sputum Xpert Ultra Semiquantitative grade | Detected by Method 2 |  | Detected by Method 3 |  |
| --- | --- | --- | --- | --- |
|  | trace+ for HIV+ only | trace+ for all | trace+ for HIV+ only | trace+ for all |
| Trace | 0/2 (0%) | 0/2 (0%) | 0/2 (0%) | 0/2 (0%) |
| Very Low | 0/6 (0%) | 0/6 (0%) | 3/9 (33.3%) | 1/5 (20.0%) |
| Low | 4/12 (33.3%) | 6/12 (50.0%) | 3/9 (33.3%) | 5/9 (55.6%) |
| Medium | 13/15 (86.7%) | 15/15 (100%) | 9/11 (81.8%) | 11/11 (100%) |
| High | 20/21 (95.2%) | 21/21 (100%) | 10/10 (100%) | 10/10 (100%) |
| All | 37/56 (66.1%) | 42/56 (75.0%) | 22/37 (59.5%) | 27/37 (73.0%) |

**Table 5.** Semiquantitative results.

|  |  | Tongue swab Xpert Ultra Method 2 |  |  |  |  | Total |
| --- | --- | --- | --- | --- | --- | --- | --- |
|  |  | Negative | Trace | Very low | Low | Medium |  |
| Sputum Xpert Ultra* | Negative | 127 | 0 | 0 | 0 | 0 | 127 |
|  | Trace | 2 | 0 | 0 | 0 | 0 | 2 |
|  | Very low | 6 | 0 | 0 | 0 | 0 | 6 |
|  | Low | 6 | 3 | 0 | 3 | 0 | 12 |
|  | Medium | 0 | 3 | 5 | 7 | 0 | 15 |
|  | High | 0 | 1 | 4 | 14 | 2 | 21 |
| Total |  | 141 | 7 | 9 | 24 | 2 | 183 |

\*Highest semiquantitative grade from sputum Xpert Ultra; test repeated if the first result = trace or indeterminate

In a subset of 37 patients, 35 (94.6%) with sputum Xpert Ultra positive (semiquantitative result of very low or higher) and 2 with sputum Xpert Ultra trace positive results, frozen tongue swab samples were also processed and tested using Method 3 (boil method). The two participants with sputum Xpert Ultra trace positive results were not detected by either tongue swab method. Among the remaining 35 participants with very low or higher semi-quantitative sputum Xpert Ultra positive results, 31 were identified by both swab methods, 2 were identified by Method 3 and missed by Method 2, and 2 were identified by Method 2 but missed by Method 3, resulting in no difference in sensitivity. When all sputum Xpert Ultra trace positive results were considered to be true positive, Method 3 identified 3 positives missed by Method 2, while Method 2 identified 1 positive that was missed by Method 3 (sensitivity difference 5.7% [95% CI -19.6 to 8.2, p-value=0.6]).

## 4. Discussion

OSA is a simpler alternative to sputum collection for TB testing, but standardized methods for pairing tongue swabs with widely available molecular testing platforms are needed. In laboratory spiking experiments, we showed that LoD is comparable to manual DNA extraction and qPCR when two tongue swabs are processed for Xpert testing using a protocol similar to that used for sputum-based testing. In clinical evaluation, this method resulted in three-quarters of patients with TB being detected by Xpert testing of tongue swabs while retaining high specificity (100%, 95% CI 95.8-100). These data highlight that OSA should be considered an alternative when sputum cannot be collected, and the need for continued optimization of OSA-based TB diagnosis to detect pauci-bacillary disease.

Compared with previous studies of OSA-based Xpert testing, we show a marked improvement in sensitivity while retaining high specificity. Sensitivity was 45% in a study from Peru that collected a single cheek swab using one of three swab types and storage buffers, and processed swabs for Xpert testing in a manger similar to Methods 1 and 2 used in our study. Based on our previous studies, more bacterial biomass is present on the tongue dorsum than on cheeks, and FLOQswabs pick up more biomass than the swab types used in the Peru study. In a study of mass TB screening among prisoners in Brazil, sensitivity was 51% overall when testing two tongue swabs sequentially and ranged from 38% to 90% in patients with very low/low to high semi-quantitative sputum Xpert results. Sensitivity was modestly higher in our study, especially among patients with higher mycobacterial load, supporting the use of our protocol involving the use of FLOQswabs and simultaneous testing of two swabs processed with Xpert SR. However, our results showed the same pattern of inability to detect paucibacillary disease.

Although our data support the use of tongue swabs when sputum cannot be collected, there remains a need to further optimize OSA-based molecular testing for TB. Our laboratory experiments showed that boiling a single swab results in lower LoD compared with processing one or two swabs with Xpert SR. However, benefits were marginal in our small clinical sub-study of the boil method. We reported recently that flocked tongue swabs collect only a small percentage of the biomass that is present on tongue dorsa [13]. Therefore, efforts to optimize sensitivity are focusing on the use of alternative swab products that collect more biomass.

Key strengths of the study include the use of laboratory- and clinic-based studies to validate protocols for Xpert testing using tongue swabs and clinical validation among a consecutive sample of patients with cough identified through clinic-based active TB case finding. However, there are also some limitations to this study. Our clinical study population is reflective of patients seeking healthcare in Uganda. OSA-based Xpert testing may have lower sensitivity if applied for TB screening among high-risk but asymptomatic individuals. Our clinical sub-study of the boil method was limited to a small subset of patients. A larger evaluation may confirm some benefits, but unless the difference is meaningful, the additional laboratory work required is unlikely to be taken up in routine practice. Last, we did not confirm positive sputum Xpert Ultra results with culture. However, Xpert has been shown to have very high specificity (>98%) for TB [20].

In summary, we demonstrate that Xpert testing of tongue swabs can be a viable alternative when sputum collection is not feasible and should be done using standardized protocols. With further improvements in sample collection and processing and leveraging advances in molecular testing of swabs for SARS-CoV-2, OSA-based molecular testing has the potential to open the door to expanded TB case-finding efforts. This non-invasive, fast, and safer diagnostic specimen and processing method may provide a useful new tool in the global fight against TB.

## Data Availability

All data produced in the present study are available upon reasonable request to the authors

## Acknowledgements

This research was supported by NIH (NIAID) grant #s U01AI152087 (AC) and R01AI139254 (GC), and Global Health Labs (AC). We are grateful to Cepheid, Inc., for providing research-use-only Xpert Ultra test kits for LoD experiments, to Copan Italia for providing swabs, and to the patients and staff at Mulago National Referral Hospital and Kisenyi Health Center IV.

