## Supplementary material for "Accuracy of tongue swab testing using Xpert MTB-RIF Ultra for tuberculosis diagnosis": Table S1

**SUPPLEMENTAL INFORMATION**

**Tongue swab sample processing methods for Xpert Ultra, and determination of limits of detection (LoDs)**

**Sample Collection, Spiking, and Storage**. Tongue swab samples to be analyzed by Xpert Ultra were collected using COPAN FLOQSwabs® Regular Flocked Swab with an 80 mm or 30 mm breakpoint (COPAN Diagnostics Inc.). These swabs were shown to maximize tongue dorsum biomass collection, relative to other products [1]. Participants self-swabbed along the breadth of the mid-tongue dorsum, firmly pressing and rolling the swab head for approximately 10 seconds. Swab heads were then immediately spiked with 10 µL of serially-diluted cultured *Mycobacterium tuberculosis* (MTB) strain H37Ra stored in 1x phosphate buffered saline with 15% glycerol and 0.05% Tween® 80 (PBSGT), or with blank PBSGT. Swab heads were then broken off into 5 mL polypropylene transport tubes (Corning) containing 800µL sterile 1x Tris-EDTA (TE) buffer (10 mM Tris-HCl containing 1 mM EDTA•Na_2_, pH 8.0) (Corning) for Methods 1-3. In Method 1 (1 FLOQSwab, Boil) and Method 2 (1 FLOQSwab, SR), each sample tube contained just one swab head, whereas in Method 3 (2 FLOQSwabs, SR), each sample tube contained two consecutively collected swab heads inserted side-by-side. Samples were either frozen at -80 °C until the day of processing or were processed the same day of collection.

Tongue swab samples to be extracted using the manual QIAGEN QIAamp DNA mini kit spin column protocol and analyzed by qPCR were collected as described previously [2] except that tongue swabs were self-collected using COPAN FLOQSwabs as described above, rather than by study personnel using alternative swab products. Swab heads were spiked with MTB H37Ra as above, then broken off into 2 mL Fisherbrand™ microcentrifuge tubes with screw caps (Thermo Fisher Scientific) containing 500µL sterile 1x TE buffer. In this method, each sample tube contains just one swab head. Samples were frozen at −80°C until the day of processing.

**Methods 1 and 2: Single (Method 1) and Double (Method 2) FLOQSwab, SR**. Samples were thawed at ambient temperature for a minimum of 30 minutes, then processed under biosafety cabinet. Two volumes (1600µL) of Cepheid Sample Reagent (SR) were added to the samples, vortexed for 10-15 seconds, and incubated for 5 minutes before vortexing for an additional 10-15 seconds. The samples were further incubated for additional 10 minutes at ambient temperature. The entire recoverable sample volume (2.0-2.4 mL) was then drawn up into a sterile transfer pipette (included in the Xpert Ultra kit) and dispensed into the sample reservoir of the Xpert Ultra cartridge, which was then loaded into a GeneXpert module, and run in. Samples were recorded as positive for MTB if GeneXpert software returned any positive result, including “Trace” diagnoses, in which the IS1081/6110 probe was positive but the rpoB probes were not.

**Sample processing Method 3: Single FLOQSwab, Boil (Boil method)**. Samples were thawed or held at ambient temperature for a minimum of 30 minutes prior to processing, and heated to 100°C for 10 minutes using a dry block heater (Thermo Scientific™ Digital Dry Baths/Block Heater) with water added to the block wells. Samples were then removed from the heat block and allowed to cool either on ice or at ambient temperature for 5 minutes (rationale and comparison of these two methods can be found in the Supplemental Material; no significant difference was observed between the two). After cooling, two volumes (total 1600µL) of sterile 1x TE buffer were added to the samples, for a total sample volume of 2.4 mL. The samples were shaken on a lab vortexer (GENIE® SI-0236 Vortex-Genie 2 Mixer, 120V) on setting 10 for 10-15 seconds, then allowed to sit at ambient temperature for 5 minutes and then shaken for an additional 10-15 seconds, before allowing them sit for additional 10 minutes before 2mL was drawn up in the sterile transfer pipette for cartridge inoculation and analysis as described below for Methods 1 and 2.

**Manual Extraction and qPCR Analysis.** DNA extraction and concentration were accomplished using the modified QIAGEN QIAamp DNA mini kit (#51306) spin column protocol and an ethanol precipitation, as previously described (12, 13). Prior to opening the samples, they were heated to 100 °C for 10 minutes in a heat block. After the addition of Buffer AL, proteinase K, and ethanol, 700µL of sample was loaded onto the spin column, with the remainder stored at -80°C as a reserve in case retesting the sample was necessary. After wash steps and as before (13), the samples were eluted twice using 150µL of Buffer AE for a final volume of 300µL, all of which was concentrated by ethanol precipitation to maximize sensitivity. The resulting dried DNA pellet was resuspended in 5µL Buffer AE.

Master mix consisted of 1× Luna Universal Probe qPCR Master Mix (New England BioLabs, Inc., Cat # M3004L), 0.45 μM forward primer, 1.35 μM reverse primer, 0.25 μM FAM/MGBNFQ probe, 2.375μL H2O. Each reaction comprised of 20µL master mix, which was added directly to the resuspended DNA pellet. After an additional 10-minute incubation, during which the samples were vortexed at medium speed to mix, the total 25µL volume was transferred to the PCR plate. Quantitative PCR was performed on the Applied Biosystems StepOnePlus Real-Time PCR system using the following reaction protocol: initial incubation at 95°C for 10 min and 45 cycles of 95°C for 15 seconds (denaturation) and 60°C for 1 minute (annealing/extension). The primers, targeting IS6110 are those designed and described previously (20, 21). Samples were recorded as positive for MTB if the Cq value was ≤ 38, as was used in previous clinical evaluation (13).

**Determination of LoDs**. LoD’s of the experimental methods were quantified using volunteer tongue swabs spiked with serial dilutions of cultured MTB H37Ra. Raw dose-response results are presented in Table S1. Exploratory runs at some dilutions (e.g. 100 cfu/swab) increased the number of runs at those dilutions, relative to other dilutions. Results of all experiments were used in LoD calculations. This minimized bias that could have resulted from the designation of specific runs as exploratory.

RStudio (Version 1.4.1103, RStudio, PBC) was used to model LoD plots and for inferential statistics. LoD plots were modelled using modified code developed by Weir et al (22). This modelling program used our experimental dose-response data to fit an exponential model and obtain Maximum Likelihood Estimation (MLE) parameters. Exponential models passed a goodness of fit test based on deviances from the MLE using a Chi-Squared distribution with an alpha of 0.05. Models were subjected to a bootstrap routine over 10,000 iterations to determine confidence intervals. Modifications of the original code included changing ID50/LD50 references and calculations to 95% LoD references and calculations. Sensitivity and LoDs are reported on a colony forming unit (CFU) per swab basis (which is the same as the CFU per sample amount for Methods 1, 2, and QIAGEN, but is half of the CFU per sample basis for Method 3 which includes 2 swabs). LoD plots are shown in Figure S1.

**Table S1. Dose response data used to plot the LoDs**

|  | **CFU/Swab** | | | | | | | | | | |
| --- | --- | --- | --- | --- | --- | --- | --- | --- | --- | --- | --- |
|  | **5** | **10** | **15** | **25** | **37.5** | **50** | **100** | **125** | **250** | **500** | **1000** |
| **Method 1** | 5/14* | 11/13 |  |  |  |  | 8/8 |  |  |  | 3/3 |
| **Method 2** | 1/5 | 8/21 | 1/4 | 1/2 |  | 3/4 | 18/20 | 3/3 | 3/3 | 2/2 | 5/5 |
| **Method 3** | 1/7 | 3/10 | 4/9 | 6/9 | 4/5 | 6/7 | 10/10 | 7/7 |  |  |  |
| **qPCR** | 1/10 | 4/10 |  |  |  | 10/10 | 10/10 |  |  |  |  |

*****Number of samples with positive result/number of samples tested

**Figure S1. Limit of detection (LoD) plots for Methods 1 through 3 and manual qPCR method.** A, Method 3; B, Method 1; C, Method 2; D, manual qPCR. A brief description of each methods appears at the top of the each panel. SR, Cepheid Sample Reagent.

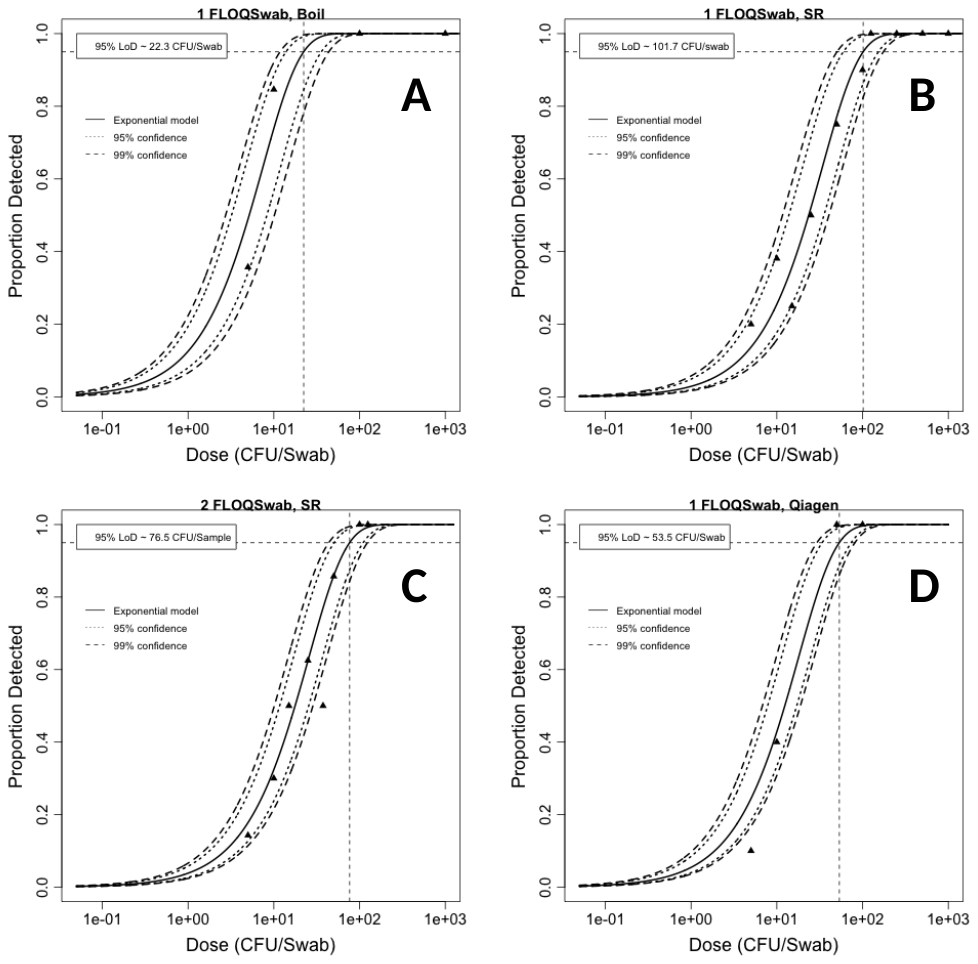
